# Point-of-Care Air Surveillance of Respiratory Pathogens Using the GeneXpert® System

**DOI:** 10.64898/2026.06.23.26354644

**Authors:** Barikisu A. Ibrahim, Tola Ewers, Isla E. Emmen, Maggie Kester, Amy L. Ellis, Jacob Meuler, Olivia Duval, Emma Copen, Mojgan Golzy, Caitlyn Kurtz, Ari N. Machtinger, Chris Crnich, David H. O’Connor, Marc C. Johnson, Shelby L. O’Connor

**Author notes:** Address correspondence to Shelby L. O’Connor,.

## Abstract

Advances in air-based surveillance of pathogen genetic material are hindered by reliance on centralized, time-consuming molecular techniques. Point-of-care (POC) diagnostic platforms, like the Cepheid® GeneXpert®, offer rapid, simplified testing in clinical settings but have not been evaluated for use with air samples. Here, we paired the ThermoFisher™ AerosolSense™ air sampler with the Xpert® Xpress SARS-CoV-2/Flu/RSV Plus test to evaluate near-real-time air surveillance. To assess analytical sensitivity, we spiked collection substrates with inactivated viruses and performed overnight sampling using the air sampler. As few as 10 copies of influenza A/B (IAV/IBV) and RSV applied to the substrate were detectable by GeneXpert, while SARS-CoV-2 required at least 100 copies for detection. Longitudinal air surveillance was conducted across congregate settings in Columbia, Missouri, and Madison, Wisconsin, in 2024-2025, collecting 281 air samples. SARS-CoV-2 was detected most frequently, followed by IAV. To assess concordance, 191 samples with paired GeneXpert and RT-qPCR results were analyzed across multiple Ct value cutoffs. Agreement between GeneXpert and RT-qPCR for SARS-CoV-2 was fair to moderate (κ = 0.306–0.443). Optimal GeneXpert Ct cutoffs for the best balance between sensitivity and specificity, determined using analyses such as Youden’s index, were site-specific: 45 for Wisconsin (67% sensitivity, 83% specificity) and 41 for Missouri (76% sensitivity, 62% specificity), reflecting differences in laboratory protocols. For IAV, agreement was moderate (κ = 0.56) with GeneXpert Ct cutoff of 40, achieving 85% sensitivity and 81% specificity. Further studies across diverse settings and viral targets are needed to establish GeneXpert’s role in routine air surveillance.

**Importance:** This study demonstrates that point-of-care molecular testing with the Cepheid® GeneXpert® provides a faster alternative to the time-consuming molecular workflows currently used for detecting airborne respiratory pathogens. The GeneXpert provides results within 37 minutes after sample addition to the Xpress CoV-2/Flu/RSV test cartridge (following approximately 20 minutes for sample elution), compared to the multiple hours or days required by laboratory-based methods, enabling mitigation measures before widespread transmission occurs. Integrating GeneXpert testing with air sampling offers a simple, semi-automated approach for pathogen detection in healthcare and community settings. Because GeneXpert instruments are already deployed in over 180 countries, including clinics in low- and middle-income settings, this approach can extend air sampling–based surveillance to regions where individual patient testing is not feasible. These workflows can also be adapted to detect other pathogens with point-of-care assays.

## 1. Introduction

Respiratory viruses such as severe acute respiratory syndrome coronavirus 2 (SARS-CoV-2), influenza A/B, and respiratory syncytial virus (RSV) contribute significantly to seasonal hospitalizations and public health burden (1–3). SARS-CoV-2 has caused over 7 million deaths globally, while seasonal influenza and RSV account for hundreds of thousands of additional respiratory deaths annually (3–6).

Environmental surveillance tools, particularly wastewater monitoring, have expanded to track community-level viral transmission (7,8). While effective for large populations, wastewater surveillance lacks spatial resolution due to its broad community-based coverage and is constrained to areas with centralized sewage infrastructure. Air sampling offers a complementary strategy for detecting airborne pathogens in indoor spaces (9–11), providing greater flexibility for targeted monitoring in high-risk congregate settings such as healthcare facilities, schools, and transit hubs.

Air sampling has shown utility for monitoring SARS-CoV-2 genetic material in hospitals and public buildings (12–14), but existing detection methods have significant limitations. Traditional viral culture methods are complex, time-consuming, and often impractical for routine surveillance due to the need for specialized cell culture facilities, extended incubation periods of 7-10 days, and frequent failure to recover viable virus from air samples (15). Laboratory-based molecular methods, such as quantitative polymerase chain reaction (qPCR) (16–18), require specialized personnel and sophisticated laboratory equipment, necessitating sample transport to centralized facilities and creating delays that limit rapid deployment.

Point-of-care molecular testing could address these limitations by enabling rapid, on-site detection with simplified protocols, particularly critical in congregate settings such as nursing homes with vulnerable populations, where prompt pathogen identification can implement control measures to reduce transmission of pathogens (19). Several advances have been made toward this goal, such as the pathogen Air Quality (pAQ) monitor, which has been shown to detect SARS-CoV-2 in the air within minutes (20), but this system is highly specialized and not commercially available. The GeneXpert® system is a rapid, automated cartridge-based nucleic acid amplification and detection system in use in more than 180 countries and supported by both commercial channels and a global access program that subsidizes deployment in high-disease-burden regions (21–23). The GeneXpert system has been successfully applied to detect pathogens in wastewater (24–27); however, it has not been evaluated for the detection of pathogen genetic material from air samples.

In this study, we evaluated the Xpert® Xpress CoV-2/Flu/RSV Plus test for rapid detection of genetic material respiratory viruses collected from the air on the GeneXpert system. This 37-minute multiplex test detects SARS-CoV-2, influenza A, influenza B, and RSV using a streamlined workflow that requires only adding the sample to a single-use cartridge for automated extraction and RT-qPCR amplification of viral RNA. By combining air sampling with this globally accessible GeneXpert system, we demonstrate a practical approach for expanding rapid respiratory pathogen surveillance.

## 2. Materials and Methods

### 2.1 Serial dilutions of inactivated viruses to be detected using GeneXpert

Inactivated SARS-CoV-2 (BEI Resources Cat No. NR-52286), IAV (H1N1) (BEI Resources Cat No. NR-49450), RSV (Microbiologics Cat No. HE0028N), and IBV (BEI Resources Cat No. NR-18924) were obtained, and stock concentrations were quantified using RT-qPCR (details for RT-qPCR assays can be found under the Detection of SARS-CoV-2, IAV, and Human RNase P by RT-qPCR section below). Serial dilutions of each inactivated virus were then prepared in a Biosafety Cabinet, and 10 μL of each dilution was applied to one clean ThermoFisher™ AerosolSense™ substrate made of polyurethane foam. The substrate was allowed to dry for 1 hour, then placed in clean AerosolSense cartridges using sterile forceps. Cartridges were then run on an AerosolSense air sampler for 24 hours in an empty, closed room (to ensure only the spiked viruses were captured). The AerosolSense pulls in air continuously at a flow rate of 200 liters per minute. The following day, the substrate was submerged in 500 μL of phosphate-buffered saline (PBS) for approximately 10 minutes at room temperature, then squeezed to release the trapped particles into the PBS. The required volume of eluate (300 μL) per the Xpert Xpress CoV-2/Flu/RSV Plus test instructions was dispensed into a Xpert Xpress CoV-2/Flu/RSV *plus* test cartridge (Cepheid, Sunnyvale, CA, USA), then inserted into the GeneXpert instrument and run using the Xpress CoV-2/Flu/RSV *plus* assay.

### 2.2 Collection of air samples from congregate spaces in Madison, WI and Columbia, MO

Two ThermoFisher AerosolSense air samplers were placed in a long-term care facility in Madison, WI. The first air sampler ran from July 2024 to April 2025, whereas the second ran from October 2024 to February 2025. The cartridges were inserted into the air samplers and then run for 1-7 days each (specific run periods for all sampling locations are listed in Supplementary Table 2). In Columbia, MO, one air sampler was placed in the University of Missouri-MU Student Center, and the second one in the Boone County Courthouse. The air sampler in the Student Center ran from October 2024 to March 2025, whereas the one in the Boone County Courthouse ran from December 2024 to March 2025. All air samplers were placed on high-top counters (above 3m high from the floor) in areas with sufficient human traffic (Figure 2). The cartridges were inserted into the air samplers and then run for 1-7 days each at both locations; however, on some days, the sampling periods exceeded 2 weeks due to certain conditions (specific run periods for all sampling locations are listed in Supplementary Table 2).

### 2.3 Detection of pathogens captured on AerosolSense substrates using GeneXpert Xpert Xpress CoV-2/Flu/RSV Plus test

Two AerosolSense substrates were removed from an air sampler cartridge, and each substrate was transferred to a 2 mL tube containing either 500 μL of phosphate-buffered saline (PBS) in the Wisconsin lab or 500 μL of Dulbecco’s Phosphate Buffered Saline (DPBS) in the Missouri lab, fully saturating the substrates for 20 minutes prior to removing the sponge and squeezing out absorbed DPBS. We note that airflow patterns within the sampler may result in an unequal distribution of material across the two substrates. Eluate (300 μL) from the first substrate was dispensed into an Xpert Xpress CoV-2-/Flu/RSV *plus* cartridge (Cepheid, Sunnyvale, CA, USA) per manufacturer’s instructions and inserted into the GeneXpert instrument, then run using the Xpress CoV-2/Flu/RSV *plus* assay.

### 2.4 RNA extraction for laboratory-based RT-qPCR detection of SARS-CoV-2, IAV, and Human RNase P

#### RNA Extraction Protocol from Madison

The second air sample substrate was eluted upon arrival at the laboratory using the same elution protocol described above. RNA was extracted from 300 μL of eluate using the Promega Maxwell RSC viral total nucleic acid kit (Promega, Madison, WI, USA) on a Maxwell 48 instrument, with an elution volume of 50 μL. This was done following the manufacturer’s protocol. The resulting RNA samples were stored at −80°C until later use.

#### RNA Extraction Protocol from Columbia

For each air sample eluate, 700 μL Viral Lysis Buffer and 5.6 μL carrier RNA (cRNA) were combined, vortexed, and 560 μL of the mixture was transferred to a 2 mL screw-cap tube without a skirted base. Air sample eluate (140 μL) was added to each 2 mL screw-cap tube for a total volume of 700 μL. This lysate was subjected to RNA extraction using the QIAamp DSP Viral RNA Mini Kit (Qiagen, Germantown, MD, USA) per the manufacturer’s instructions, using the QIAGEN QIAcube Connect and the QIAcube QIAamp Viral RNA Manual lysis program, with an elution volume of 50 μL. The resulting RNA samples were stored at −20°C until later use.

### 2.5 Detection of SARS-CoV-2, IAV, and Human RNase P by RT-qPCR

#### RT-qPCR protocol for quantifying inactivated virus stocks in Madison

The number of copies of viral RNA in the stock inactivated viruses was quantified by RT-qPCR on a LightCycler480 (Roche) using the TaqMan Fast Virus 1-step qPCR master mix (ThermoFisher Cat No. 5555532) and primers/probes targeting each virus listed in Table 1. SARS-CoV-2 primer/probe mixes were obtained from IDT (Coralville, IA, USA); IAV, IBV, and RSV primers were made in-house. Cycling conditions were as follows: for SARS-CoV-2, 37 °C for 2 min, 50 °C for 15 min, and 95 °C for 2 min, followed by 50 cycles of 95 °C for 3 seconds and 55 °C for 30 s, then cooling for 30 s at 37°C; for IAV, 50 °C for 10 min and 95 °C for 20 s, followed by 50 cycles of 95 °C for 15 s and 62 °C for 1 min, then cooling for 30 seconds at 40 °C; and for IBV and RSV, 50 °C for 5 min and 95 °C for 20 s, followed by 50 cycles of 95 °C for 15 s and 60 °C for 90 s.

**Table 1:**
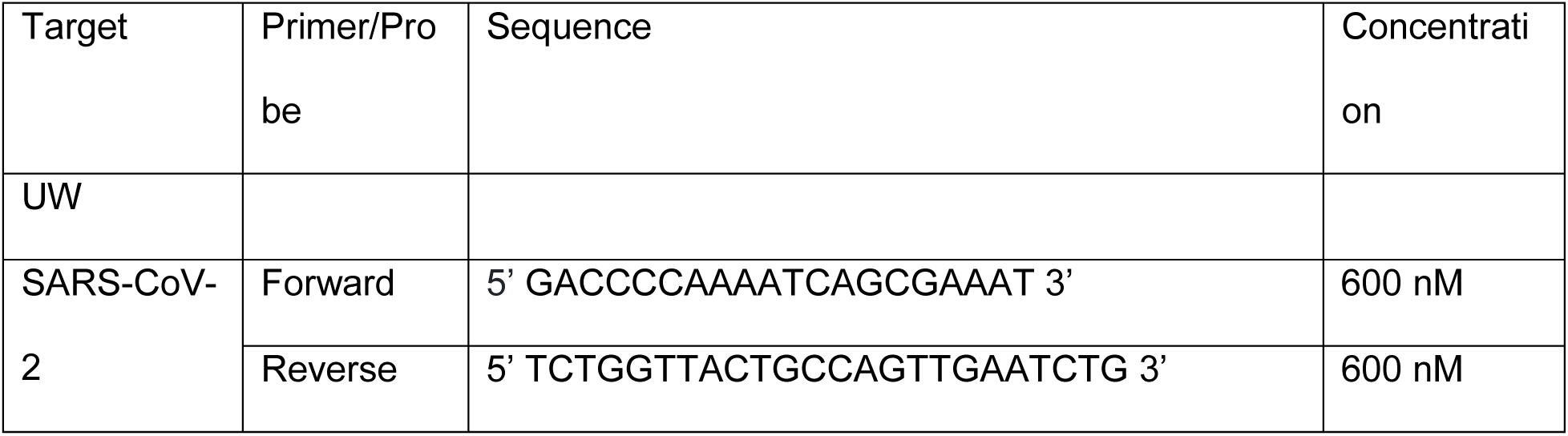
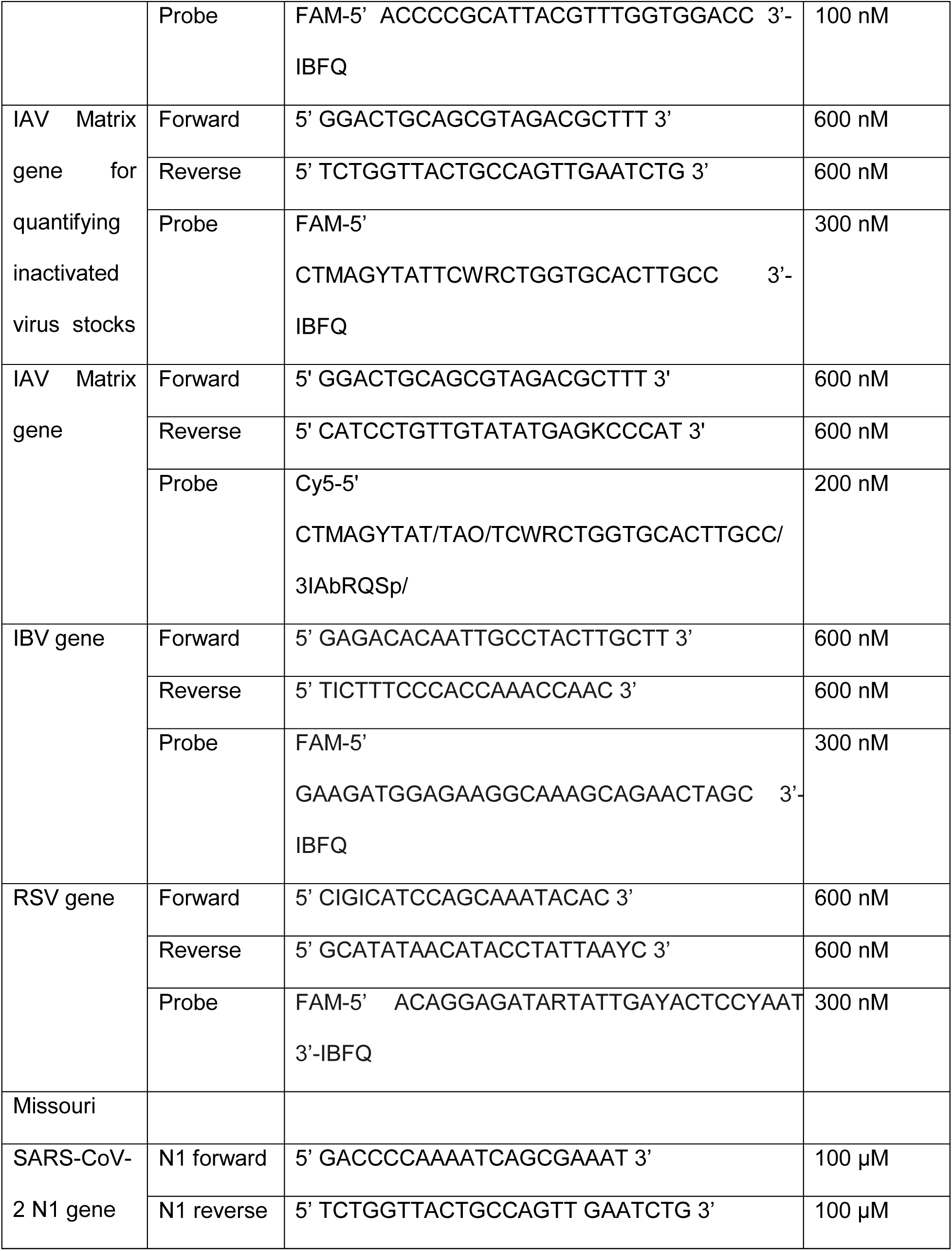
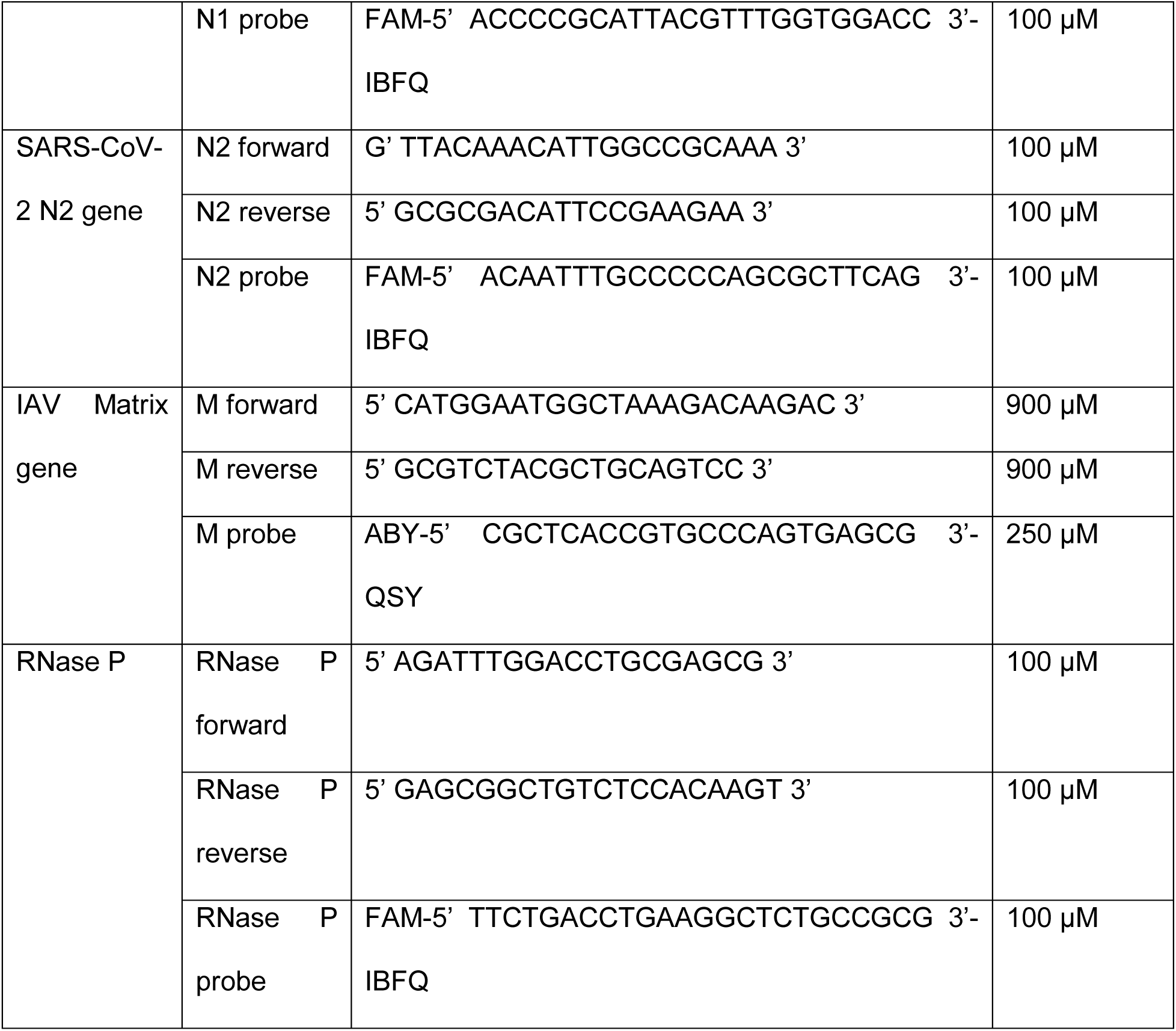
Primers and probes for SARS-CoV-2, IAV, IBV, RSV, and RNase P.

#### RT-qPCR protocol for air samples in Madison

RT-qPCR was performed on RNA extracted from air samples collected from the long-term care facility to detect and quantify SARS-CoV-2 and IAV using a multiplex PCR assay. The 20 µL RT-qPCR reaction mix consisted of nuclease-free water, TaqMan Fast Virus 1-Step (enzyme), forward/reverse primers combined with random hexamers, probes (N1-FAM and M-Cy5), and 10 µL of the isolated RNA. SARS-CoV-2 primer/probe mixes were obtained from IDT (Coralville, IA, USA) and IAV primers were made in-house. Human RNase P, as an endogenous control for human genetic material in each sample, was tested in a separate reaction. This RT-qPCR reaction mix consisted of nuclease-free water, TaqMan Fast Virus 1-Step (enzyme), the RNase P primer/probe mix prepared by IDT (Coralville, IA, USA), and 5 μL of isolated RNA. Cycling conditions for all targets were as follows: 50°C for 10 min, 95°C for 20 sec, followed by 50 cycles of 95°C for 15 sec and 62°C for 1 min, and a final cool step at 40°C for 30 sec (primers and probes are listed in Table 1). For all RT-qPCR assays, viral RNA copies per reaction were calculated based on a standard curve.

#### RT-qPCR protocol for air samples in Columbia

Each RT-qPCR reaction consisted of nuclease-free water, TaqPath 1-Step Multiplex Mastermix (ThermoFisher Cat No. A15299), primer/probe mix (primers are listed in Table 1), and 5 µL of RNA extract were added to an Applied Biosystems MicroAmp 96-well reaction plate (Cat No: 4346907). The plate containing 20 μL in each well was spun down and run on the Applied Biosystems 7500 Fast System. Cycling conditions for the detection of both IAV and SARS-CoV-2 were as follows: 25°C for 2 min, 50°C for 15 min, 95°C for 2 min, followed by 45 cycles of 95°C for 15 sec, and then a final step of 60°C for 1 min. Both viruses were detected using either a single or a multiplex PCR assay (primers and probes are listed in Table 1). SARS-CoV-2 primer/probe mixes were obtained from IDT (Coralville, IA, USA) and IAV primers were made in-house. Human RNase P was used as an endogenous control for human genetic material in each sample.

### 2.6 Comparing GeneXpert and RT-qPCR Results

#### Statistical Analysis

To evaluate diagnostic agreement between GeneXpert and RT-qPCR, RT-qPCR was designated as the reference standard, with all detections classified as positive regardless of Ct value. GeneXpert positivity was assessed across Ct value cutoffs from 37 to 45 to determine optimal concordance thresholds. Ct values below the cutoff were considered positive, and above were negative. For each cutoff, confusion matrices were constructed to derive sensitivity, specificity, positive predictive value (PPV), and negative predictive value (NPV) with 95% confidence intervals. Cohen’s kappa was calculated to quantify agreement beyond chance, with standard interpretation thresholds: <0.20 (poor), 0.21–0.40 (fair), 0.41–0.60 (moderate), 0.61–0.80 (substantial), and >0.80 (almost perfect). Additional metrics included Matthews correlation coefficient (MCC) for class imbalance robustness, F1 score as the harmonic mean of sensitivity and PPV, and McNemar’s test to assess systematic bias between methods. All analyses were conducted in R (version 4.x) using irr, dplyr, readxl, and writexl packages. R codes are uploaded in the supplementary file.

To define the optimal cutoff point for classifying positive and negative air sample results on GeneXpert, we also performed Receiver Operating Characteristic (ROC) curve analysis (Supplement Figure 1). We fitted separate logistic regression models for Missouri, Wisconsin, and combined SARS-CoV-2 datasets, with RT-qPCR results as the outcome and GeneXpert Ct values as the predictor. Area Under the ROC Curve (AUC) was calculated to assess discriminatory ability, with values of 0.7–0.9 considered good and >0.9 excellent (Supplementary Table 6; Supplementary Figure 1). Optimal cutoffs were determined using two complementary approaches: 1) maximizing Youden’s Index (J = Sensitivity + Specificity - 1), which balances true positive and false negative rates, and 2) minimizing Euclidean distance to the perfect classification point (D = √[(1-sensitivity)² + (1-specificity)²]). The final optimal threshold was defined as the value that simultaneously maximized Cohen’s kappa, F1 score, MCC, and Youden index (J) while minimizing Euclidean distance.

SARS-CoV-2 analyses were performed separately for Missouri and Wisconsin sites, because they differed in RNA extraction input volumes (140 µL and 300 µL, respectively). Influenza A was evaluated using only Missouri samples, with the GeneXpert IAV A1 and IAV A2 channels analyzed independently.

## 3. Results

### 3.1.1 Analytical Sensitivity of the GeneXpert System Using Inactivated Viruses

The GeneXpert Xpert Xpress CoV-2/Flu/RSV Plus test was initially validated for clinical nasal and nasopharyngeal swabs, with limits of detection established for SARS-CoV-2 variants, IAV subtypes, IBV, and RSV (28). Here, our goal was to determine the minimum viral genome copy number required to yield a positive Ct result on the GeneXpert system when processed through our air-sampling workflow. We applied serial dilutions of inactivated viruses to air substrates, ran them for 24 hours in an empty room using a Thermo Fisher™ AerosolSense™ air sampler, and processed the cartridges using our air sampling workflow described in the methods. The eluate was tested with the GeneXpert® system using the Xpert® Xpress CoV-2/Flu/RSV Plus test. The GeneXpert system interprets results using quantitative real-time PCR cycle threshold (Ct) values and endpoint fluorescence.

We classified positivity for all viral targets based only on GeneXpert Ct values, rather than the instrument’s qualitative calls, as the assay’s interpretation algorithm was validated for clinical samples rather than air samples. SARS-CoV-2 was detected in 50% (4/8) of samples with 100 genome copies (gc) and in 100% (8/8) of samples with 1,000 gc applied to the substrate. SARS-CoV-2 was detected in 100% (4/4) of samples tested at 10,000 gc–100,000 gc. IBV was detected in 50% (2/4) of samples when 1 gc was applied and 100% (3/3) of samples at 10 gc and from 100–1,000 gc (4/4). RSV was detected in 75% (3/4) of samples when 10 gc were applied and in 100% (4/4) of samples from 100–1,000 gc. IAV was detected in 25% (1/4) of samples when 1 gc was applied to the substrate and in 100% (4/4) of samples at 10–1,000 gc. A positive IAV A1 signal indicated IAV detection (Figure 1a–d). SARS-CoV-2 required approximately 10-fold higher genome copy numbers for reliable detection than the other respiratory viruses tested, similar to Cepheid’s reported clinical detection limit of 138 copies/mL for nasal swabs (26).

**Figure 1.**
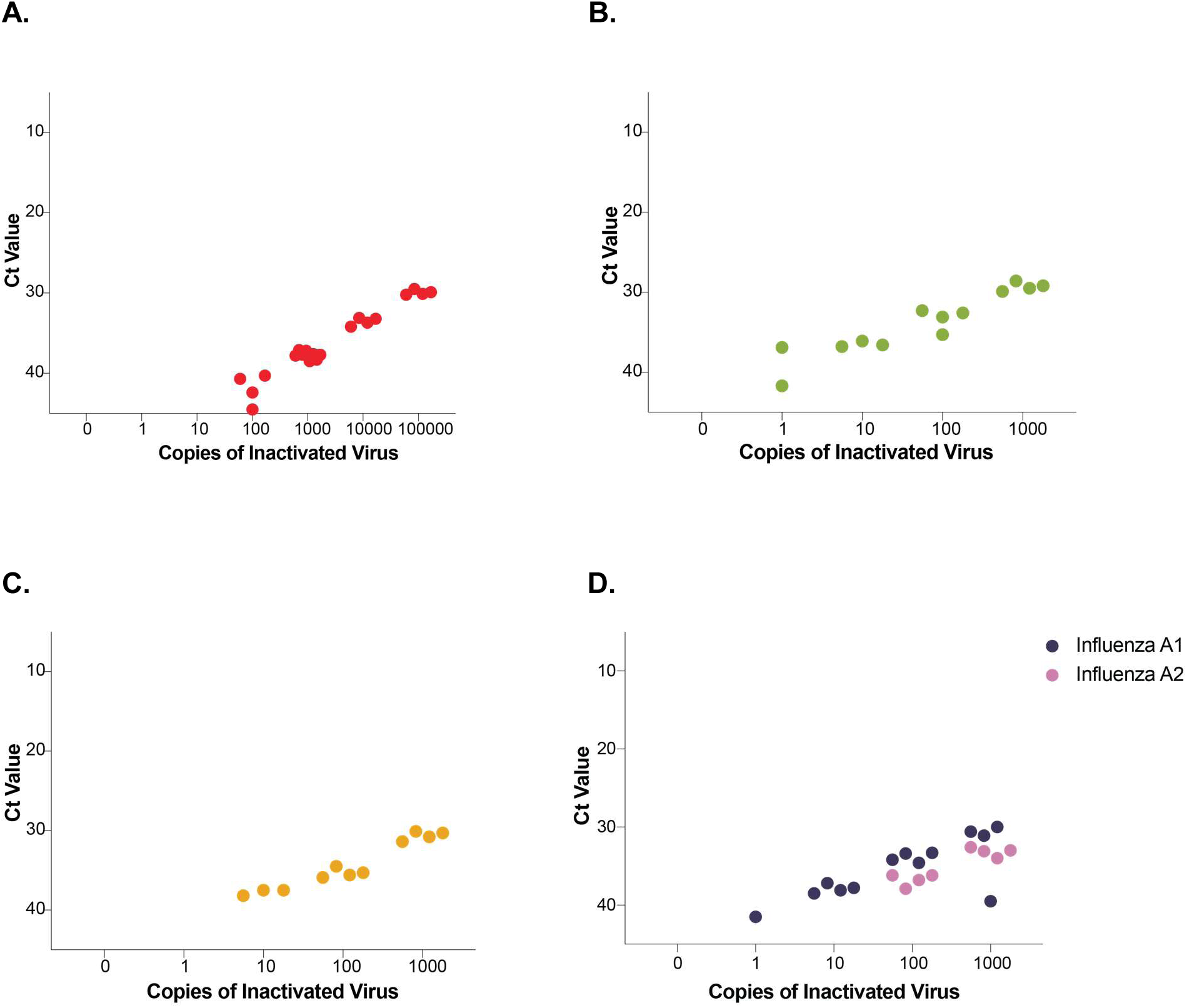
Sensitivity of the Xpert Xpress CoV-2/Flu/RSV Plus test for detecting respiratory viruses from air sampling substrates. Ten-fold serial dilutions of inactivated SARS-CoV-2, influenza B virus (IBV), RSV, and influenza A virus (IAV) (H1N1) were applied to AerosolSense air sampling substrates. Substrates were placed in cartridges and run for 24 hours on AerosolSense air samplers. Substrates were then eluted in PBS, and eluates were run on the GeneXpert platform. The x-axis shows the total copies of virus added to each substrate; the y-axis shows the Ct values from the Cepheid GeneXpert. (A-D) show the limit of detection for serially diluted copies of SARS-CoV-2 (A), IBV (B), RSV (C), and IAV A1 and A2 (D). IAV A1 and A2 are two separate channels in the Cepheid Xpert Xpress CoV-2/Flu/RSV Plus test, containing different gene targets that detect IAV strains. Positive results from either or both IAV A1 or A2 targets are considered positive IAV.

### 3.1.2 Mixed-Virus Interference Testing on the GeneXpert System

Because multiple respiratory viruses could be present in the air space simultaneously, we assessed potential interference when multiple viruses were applied to the same substrate. To test this, we co-applied 1000 gc of inactivated SARS-CoV-2 with serial dilutions of inactivated IAV (Figure 2a) or 1000 gc of inactivated IAV with serial dilutions of SARS-CoV-2 (Figure 2b). Under both co-application conditions, each virus remained detectable across all dilutions tested, with Ct values comparable to those obtained from single-virus applications, indicating no significant loss of sensitivity due to co-detection (Figure 2a-b).

**Figure 2.**
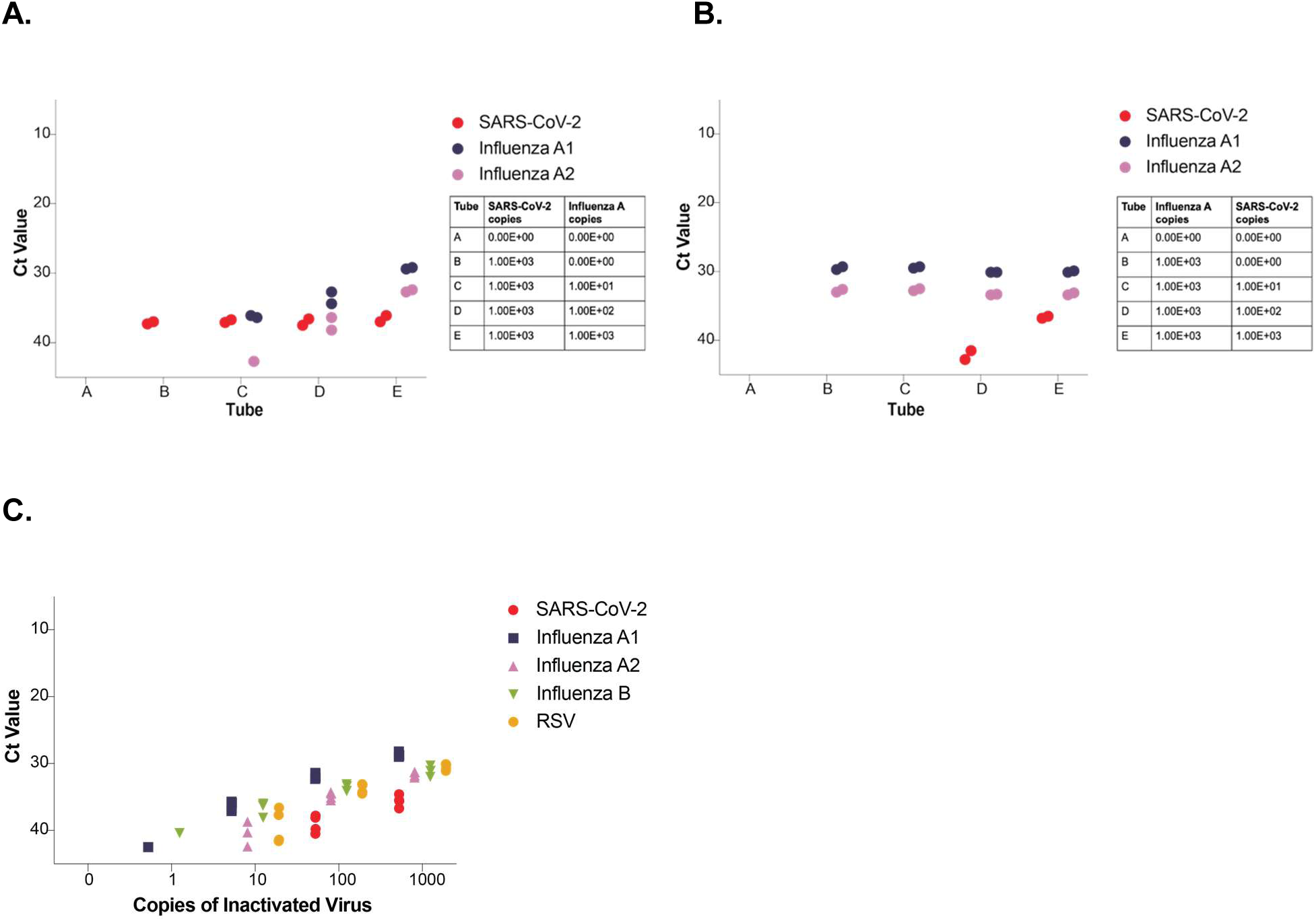
Specificity and interference testing of the Xpert Xpress CoV-2/Flu/RSV Plus test for detecting respiratory viruses from air sampling substrates. (A) A fixed amount of inactivated SARS-CoV-2 (1000 copies) along with serial dilutions of inactivated IAV were co-applied to AerosolSense substrates and run in air samplers. Subsequently, the samples were run on the GeneXpert platform, and Ct values for SARS-CoV-2 and IAV were determined. The table to the right of the graphs shows the copies of each virus applied to each substrate for Tubes A-E (x-axis). (B) A fixed amount of inactivated IAV (1000 copies), along with serial dilutions of inactivated SARS-CoV-2, were co-applied to AerosolSense substrates and run in air samplers. Subsequently, the substrates were eluted and run on the GeneXpert platform, and Ct values for SARS-CoV-2 and IAV were determined. The table to the right of the graph shows the copies of each virus applied to each substrate for Tubes A-E (x-axis). (C) shows all four viruses co-applied to the substrate at increasing copies. The x-axis shows the total copies of virus added to each substrate; the y-axis shows the Ct values from the Cepheid GeneXpert.

Lastly, we applied serial dilutions of a mix of all 4 viruses (SARS-CoV-2, IAV, IBV, and RSV) to AerosolSense substrates and performed the same workflow as above. The Ct values for each virus in the multiplex condition were not significantly different from those obtained in individual virus runs (Figure 2c; Supplementary Table 5).

### 3.2 Longitudinal Surveillance and Detection of SARS-CoV-2, IAV/B and RSV using Cepheid GeneXpert

Air sample analysis using laboratory-based RT-qPCR requires complex laboratory processing, delaying pathogen detection. To evaluate whether point-of-care molecular testing could simplify air surveillance workflows, we deployed a GeneXpert machine on-site at a congregate living health care facility in Madison, WI. We placed two AerosolSense air samplers in different locations within the facility and collected 219 air samples. Location A was sampled from July 2024 to April 2025, while location B overlapped from October 2024 to February 2025. For each collected sample, one of the two AerosolSense foam substrates was eluted and tested with the GeneXpert instrument with the Xpress CoV-2/Flu/RSV Plus assay on-site to longitudinally examine the presence of pathogen genetic material in these spaces (Figure 3). The second substrate was reserved for RT-qPCR. Genetic material for all four pathogens yielded Ct values below the detection thresholds (Ct < 45 for SARS-CoV-2, Ct < 40 for IAV, IBV, and RSV) at both locations in the facility. Using the GeneXpert platform, SARS-CoV-2 yielded positive results in 53.42% (117/219) of samples across both locations; IAV A1 and A2 targets in 15.07% (33/219) and 4.11% (9/219) of samples, respectively; IBV in 7.76% (17/219); and RSV in 2.28% (5/219) (Figure 4a-b).

**Figure 3.**
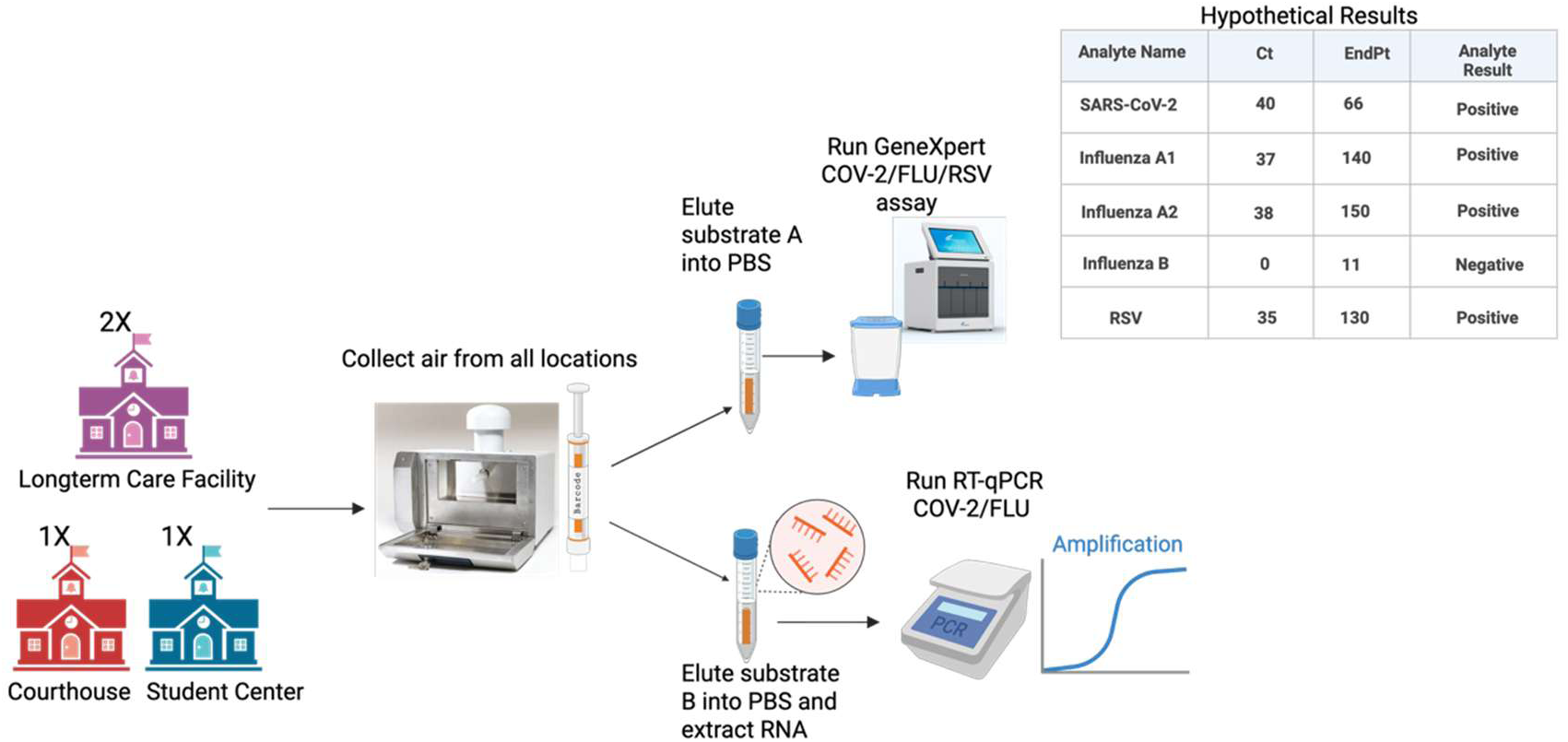
Workflow for Air Sample Collection, Processing, and Viral Detection. Air was sampled using ThermoFisher AerosolSense (image of the air sampler obtained from Thermo Fisher website) instruments at locations in Columbia, Missouri, and Madison, Wisconsin, over 24–168 hours. After collection, both substrates were eluted in PBS. One eluate from one substrate was tested directly using the Cepheid Xpert Xpress CoV-2/Flu/RSV Plus test. RNA was extracted from the second eluate/substrate, and RT-qPCR was performed as indicated in the methods to detect SARS-CoV-2 and IAV.

**Figure 4.**
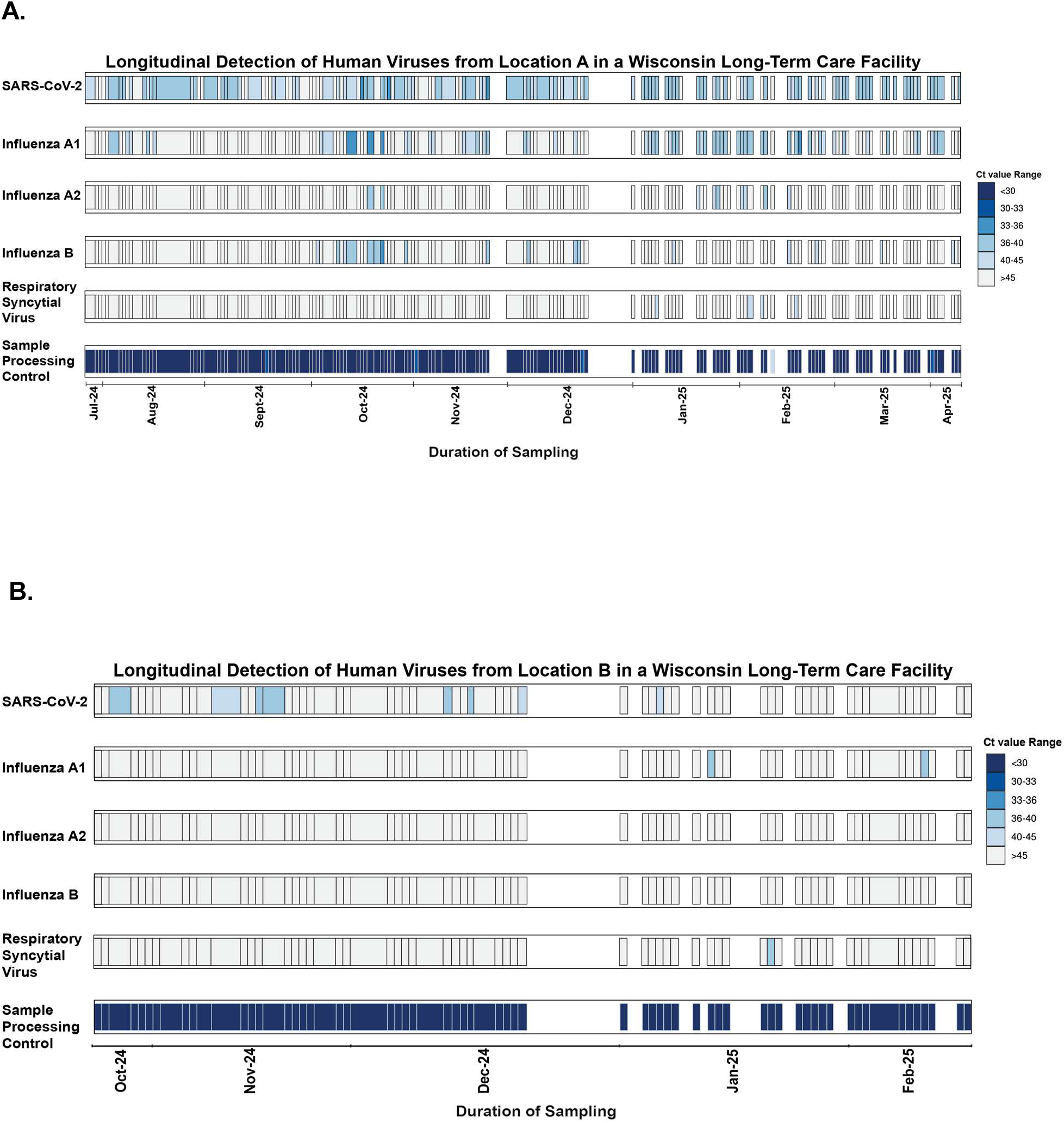
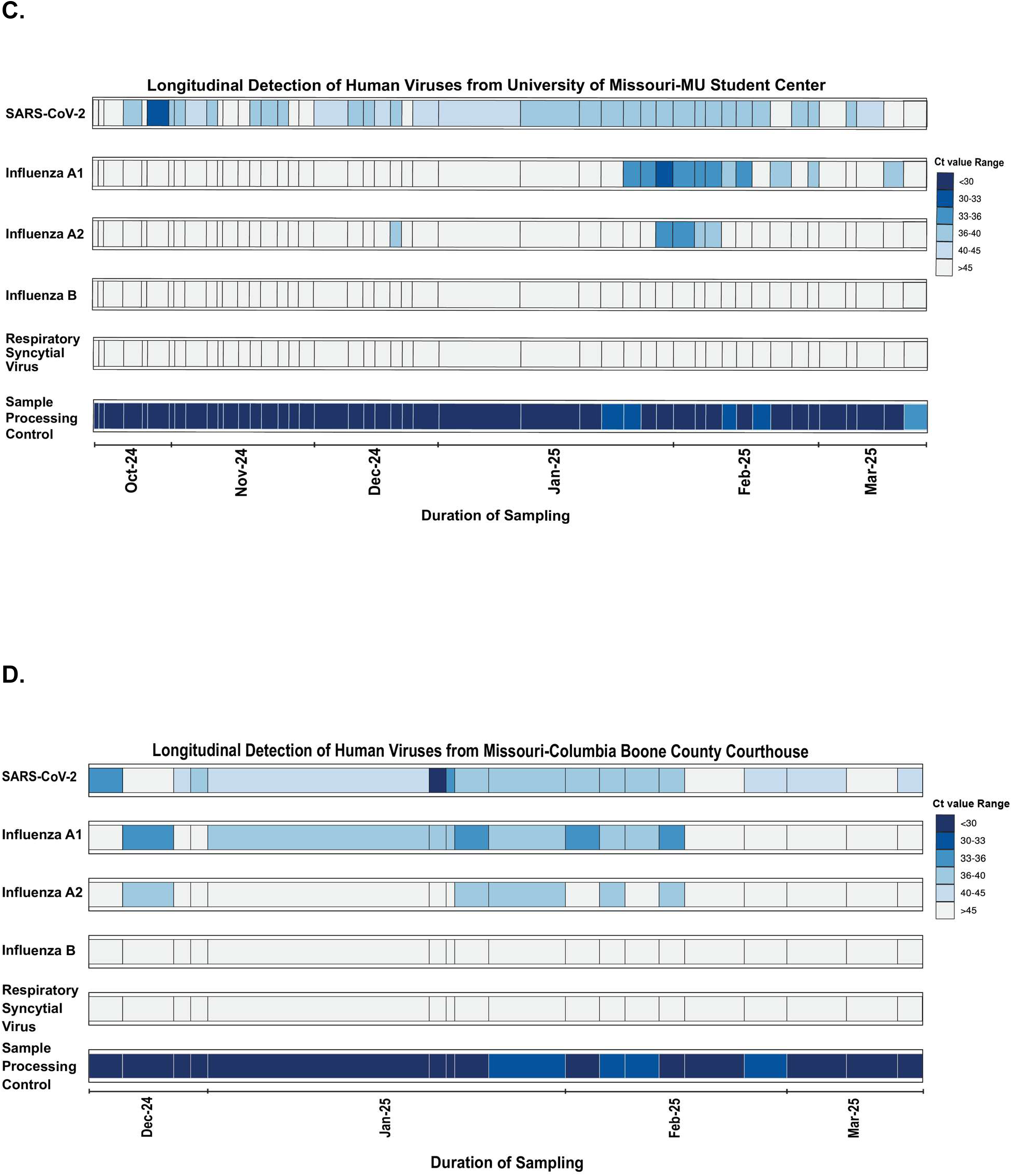
Detection of SARS-CoV-2, IAV, IBV, and RSV across two air sampling locations in Wisconsin–Madison and Missouri–Columbia. Air was sampled using AerosolSense samplers from four indoor locations across Wisconsin and Missouri between July 2024 and April 2025, and respiratory viruses were detected using the Xpert Xpress CoV-2/Flu/RSV Plus test. The x-axis indicates the sampling dates and durations. The white areas indicate periods during which no sampling was conducted. Due to longer sampling periods in January (at the Missouri Student Center and the Boone County Courthouse), the heatmaps were expanded to reflect this. However, the specific sampling period for each location is provided in Supplementary Table 2. The different pathogens and sample controls (sample processing control) are indicated on the left y-axis. The heatmap colors indicate the Ct value detected for each pathogen for each sample.

We performed a similar collection and analysis workflow at two additional locations in Columbia, MO. Location C was a University of Missouri-MU Student Center collected from October 2024 to March 2025 (Figure 4c), and Location D was a Boone County courthouse collected from December 2024 to March 2025 (Figure 4d). A total of 62 samples were collected from both locations. In Columbia, the GeneXpert instrument was housed off-site in the laboratory, but the same testing workflow used in Madison was used. SARS-CoV-2 yielded positive results in 74.19% (46/62) of samples. IAV A1 and A2 were detected in 32.26% (20/62) and 16.13% (10/62) of samples, respectively, with detections occurring between December 2024 and March 2025. IBV and RSV were not detected at either of the Columbia sites.

Across all four sampling locations, SARS-CoV-2 was consistently the most prevalent virus detected (Supplementary Table 3). IAV was the second most detected in Madison and Columbia; it was observed only between January and March, during winter in Columbia. IBV was detected only sporadically in Madison, and RSV was rarely detected and, when present, exhibited high Ct values (38< Ct<40), consistent with low viral load in the air during the sampling period.

### 3.3 Agreement between GeneXpert and laboratory-based RT-qPCR SARS-CoV-2 Ct values from air samples

We evaluated the concordance between GeneXpert and RT-qPCR for SARS-CoV-2 detection separately for each location, using air samples run on both assays: 129 samples from Wisconsin and 62 from Missouri, totaling 191 samples. RT-qPCR results were classified as positive for any detectable amplification regardless of Ct value, serving as the reference standard. To determine the optimal threshold for concordance with RT-qPCR positive/negative classifications, we tested different Ct cutoff values for defining GeneXpert positives, ranging from 37 (corresponding to 1,000 SARS-CoV-2 gc in our analytical experiments, Table 2) to 45 (the maximum cycle number, indicating detection of <100 genome copies) (Supplementary Table 6). To identify the optimal cutoff, we used multiple complementary metrics to avoid reliance on any single measure: Cohen’s kappa (agreement beyond chance), F1 score (balance of sensitivity and precision), Youden index (balance of true positive and false negative rates), and Euclidean distance to perfect classification (Supplementary Table 6; Table 3). The optimal cutoff was defined as the Ct value that simultaneously maximized Cohen’s kappa, the F1 score, and the Youden index while minimizing the Euclidean distance.

**Table 2.**
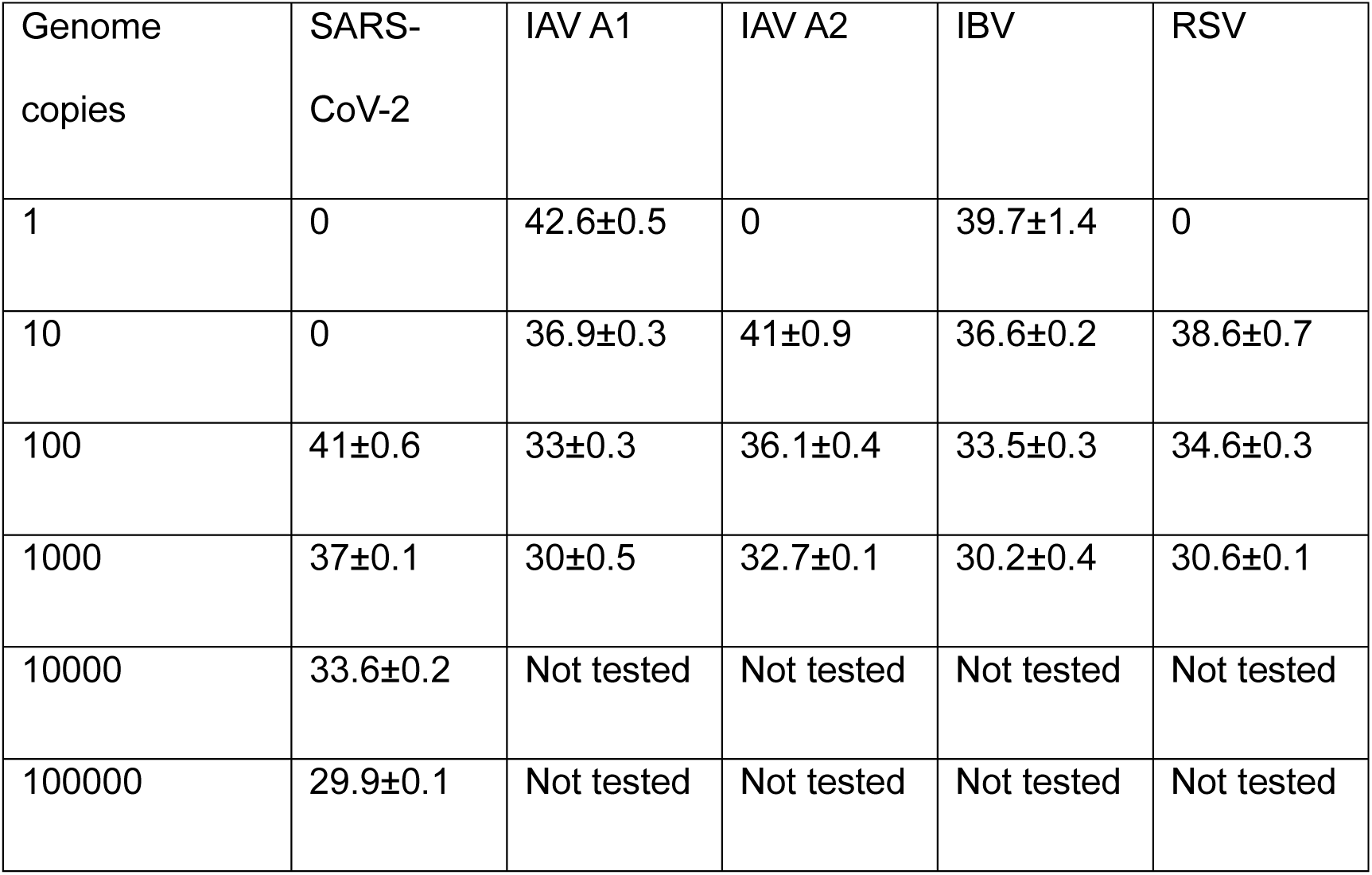
Mean GeneXpert Ct values for SARS-CoV-2, IAV, IBV, and RSV under single-virus and multiplex detection conditions.

**Table 3.** Optimal cutoff points and corresponding performance metrics for SARS-CoV-2 are presented in the following table.

| Data set | Cut-off | Sensitivity % | Specificity % | PPV % | NPV % | $\kappa$ | F1 | MCC | J | D |
| --- | --- | --- | --- | --- | --- | --- | --- | --- | --- | --- |
| Missouri | 41 | 76.3 | 62.5 | 76.3 | 62.5 | 0.39 | 0.76 | 0.39 | 0.39 | 0.44 |
| Wisconsin | 45 | 66.3 | 83.7 | 89.1 | 55.4 | 0.44 | 0.76 | 0.47 | 0.5 | 0.37 |

Using this multi-metric approach, we identified optimal cutoffs of 41 for Missouri and 45 for Wisconsin, which performed best across all evaluated metrics (Table 3). Notably, the Missouri cutoff of 41 aligns with our experimental detection threshold of 100 gc of SARS-CoV-2 in the analytical sensitivity experiments (Table 2).

### 3.4 Agreement between Cepheid and RT-qPCR IAV Ct values from air samples

We performed similar comparative analyses for IAV using the same 62 Missouri samples that were tested with both GeneXpert and RT-qPCR assays. Wisconsin samples were excluded from this concordance analysis because only 2 samples showed IAV-positive results by GeneXpert (IAV A1 and A2 targets) with Ct values below the detection threshold, which was insufficient for reliable concordance analysis, and all corresponding RT-qPCR results were negative. The same multi-metric statistical analysis used for SARS-CoV-2 was applied to determine optimal Ct cutoffs across the range of 37 to 40 (no positive detections were observed above Ct 40) (Supplementary Table 6). We identified an optimal cutoff of 40 for IAV A1, achieving the highest sensitivity, a moderate Cohen’s kappa value (k=0.57), the highest Youden index, and the shortest Euclidean distance to perfect classification (Table 4). For IAV A2, the optimal cutoff was 39, which yielded a moderate Cohen’s kappa (κ = 0.53) and the highest Youden index (Table 4). These cutoffs align with our analytical validation experiments, where IAV A1 was consistently detected below Ct 40 at 10 genome copies and above, but showed inconsistent detection above Ct 40 with 1 genome copy. Similarly, IAV A2 was consistently detected below Ct 40 at 100 genome copies and above but showed inconsistent detection above Ct 40 with 10 genome copies (and failed to detect 1 genome copy).

**Table 4.** Optimal cutoff points and corresponding performance metrics for IAV A1 and A2 are presented in the following table.

| Data set | Cut-off | Sensitivity % | Specificity % | PPV % | NPV % | $\kappa$ | F1 | MCC | J | D |
| --- | --- | --- | --- | --- | --- | --- | --- | --- | --- | --- |
| IAV A1 | 40 | 85.7 | 81.3 | 57.1 | 95.1 | 0.57 | 0.69 | 0.59 | 0.67 | 0.24 |
| IAV A2 | 39 | 50 | 95.8 | 77.8 | 86.8 | 0.53 | 0.61 | 0.54 | 0.46 | 0.5 |

## 4. Discussion

A challenge for implementing air sampling as a routine, rapid public-health surveillance tool is the long delay between collecting airborne pathogens and subsequent complex molecular testing. Standard detection workflows require nucleic acid extraction, multiple manual handling steps, and trained personnel, collectively extending processing time and limiting the feasibility of timely interventions in many settings. Here, we evaluated whether a point-of-care molecular diagnostic platform could overcome these barriers by enabling rapid, on-site testing. By eluting material from the air sample cartridge, directly adding it to the Xpert® Xpress CoV-2/Flu/RSV Plus test single-use cartridge and inserted into the GeneXpert® instrument, we avoided the complexities of separate nucleic acid isolation and molecular amplification. This approach represents a substantial improvement in operational turnaround compared to laboratory-based RT-qPCR and extends air-sampling capability to locations without access to molecular laboratories.

Our analytical sensitivity experiments demonstrate that detection of SARS-CoV-2, IAV/B, and RSV by the Xpert Xpress CoV-2/Flu/RSV Plus test on the GeneXpert system using our air sampling protocol yields a relative sensitivity similar to that obtained with the same assay on different sample types. IBV and IAV exhibited the lowest detection threshold of 1 gc, followed by RSV (10 gc) and SARS-CoV-2 (100 gc). While a direct quantitative comparison between air samples and clinical specimens is challenging due to differences in sample matrices and processing methods, relative sensitivity patterns align with clinical performance data. For SARS-CoV-2, previous studies using clinical samples reported detection limits ranging from 50-131 copies/mL with the Xpert Xpress CoV-2/Flu/RSV Plus test, compared to the manufacturer’s specification of 138 copies/mL (28,29). For influenza A and B, clinical detection limits of 50 copies/mL have been reported, while RSV shows detection at 300 copies/mL in clinical specimens(30,31). The higher sensitivity observed for IAV and IBV in our air sampling experiments is consistent with their lower clinical detection thresholds compared to SARS-CoV-2 and RSV, suggesting the assay maintains its relative sensitivity hierarchy across different sample types. Importantly, Ct values for each virus were consistent when the same genome-copy inputs were tested individually or in mixed-virus conditions, indicating that the presence of multiple respiratory viruses on a single substrate does not compromise assay performance (Table 2, Supplementary Table 5). This is particularly relevant for environmental samples, which often contain heterogeneous mixtures of viral material, cellular debris, and other biological constituents. We did not include an assessment of the elution method on the success of detection. Rather, our goal was to identify the Ct value generated by the GeneXpert instrument when a specific number of inactivated viruses were applied to the substrate and processed through a normal workflow. Alternative elution methods could further improve the detection sensitivity of pathogens collected on the cartridges. However, current technologies demonstrate that as few as 1 or 10 viral copies can be captured by an air sampler, eluted, and detected by the Xpert Xpress CoV-2/Flu/RSV Plus test on the GeneXpert. This suggests that air collection of a defined number of viral genome copies, followed by a simple elution protocol, can yield consistent Ct values with a GeneXpert.

Despite lower analytical sensitivity, SARS-CoV-2 was the most frequently detected virus across actual air samples from all sampling sites, suggesting ongoing circulation even in the post-pandemic phase. IAV was primarily detected during the winter months, indicating the virus’s seasonal dynamics (9,11,20,32). IBV and RSV were rarely detected, which was consistent with low prevalence in the community (33–36).

Agreement between GeneXpert and RT-qPCR for SARS-CoV-2 was fair to moderate across all cutoffs at both sites (Missouri: κ = 0.094–0.306; Wisconsin: κ = 0.032–0.443), with optimal cutoffs identified as 41 for Missouri and 45 for Wisconsin. The difference in optimal Ct cutoffs between Missouri (41) and Wisconsin (45) most likely reflects methodological variations between the two laboratories that affected RT-qPCR sensitivity. Wisconsin used higher input volumes for RNA extraction (300 μL versus 140 μL at Missouri) and larger template volumes for RT-qPCR (10 μL versus 5 μL at Missouri), resulting in approximately 4-fold higher viral RNA concentrations in their RT-qPCR reactions. This enhanced RT-qPCR sensitivity enabled Wisconsin to detect lower viral loads that Missouri’s protocol might miss. Interestingly, the Missouri cutoff (41) aligns with our experimental detection threshold of 100 genome copies, while Wisconsin achieved optimal performance at cutoff 45, which corresponds to detection of <100 genome copies and thus represents higher analytical sensitivity. These differences suggest that methodological variations between laboratories can impact optimal cutoff determination. For influenza A, agreement was substantially stronger, with IAV A1 achieving moderate agreement at Ct 40 while IAV A2 showed moderate to fair agreement with an optimal cutoff of 39, indicating its role as a less sensitive channel.

Unfortunately, the Influenza A concordance analysis was limited to Missouri samples. Among Wisconsin samples run on both platforms for comparison, all RT-qPCR results were negative, and only 2 GeneXpert positive results (for both IAV A1 and IAV A2) were below the detection threshold, which was insufficient for reliable analysis. Also, for influenza A, 9 samples were GeneXpert IAV A1-positive but RT-qPCR-negative, with only 2 samples showing the reverse pattern. This discordance reflects the different detection strategies employed by the assays: GeneXpert targets the matrix (M), basic polymerase (PB2), and acidic protein (PA) genes, whereas our RT-qPCR assay targets only the matrix gene. These results illustrate how assays with different gene-targeting approaches can yield different detection outcomes, an important consideration when implementing point-of-care molecular testing for air surveillance.

The findings of this study demonstrate that the GeneXpert platform can be repurposed from clinical diagnostics to environmental air surveillance, capturing the presence of SARS-CoV-2, IAV, IBV, and RSV in congregate settings without dependence on centralized laboratory infrastructure. The global deployment of more than 10,000 GeneXpert instruments, including in resource-limited settings where access to molecular laboratories is constrained, supports the scalability of this approach across diverse surveillance contexts. Beyond the pathogens evaluated here, this workflow could be adapted to other targets of public health relevance for which GeneXpert-compatible tests already exist, such as Mycobacterium tuberculosis and norovirus, enabling targeted surveillance in settings where these pathogens pose particular risks. By reducing the time between sample collection and result compared to laboratory-based RT-qPCR, this approach supports more timely public health responses to respiratory pathogen circulation. Further studies across diverse settings and viral targets are needed to fully establish GeneXpert’s role in routine air surveillance, but these findings provide a foundation for integrating point-of-care molecular testing into scalable, accessible respiratory pathogen monitoring programs.

## Acknowledgment

We thank Inkfish, LLC, for their generous support that made this work possible. We are also grateful to our collaborators and partnering institutions for providing access to sampling sites and for their continued engagement throughout this project. We acknowledge the valuable contributions of the laboratory teams in Missouri and Wisconsin for their efforts and assistance throughout the development of this study.

## Author Contribution

Barikisu A. Ibrahim, Conceptualization, Data curation, Formal analysis, Investigation, Methodology, Project administration, Validation, Visualization, Writing-original draft, Writing-review &editing | Tola Ewers, Data curation, Investigation, Methodology | Isla E. Emmen, Data curation, Investigation, Methodology, Validation, Writing-original draft, Writing-review &editing | Maggie Kester, Data curation, Methodology, Writing-original draft, Writing-review &editing | Amy L. Ellis, Data curation, Investigation, Methodology, Validation, Writing-original draft, Writing-review &editing | Jacob Meuler, Data curation, Investigation | Olivia Duval, Data curation, Investigation | Emma Copen, Investigation, Writing-review &editing | Mojgan Golzy, Formal analysis, Writing-review &editing | Caitlyn Kurtz, Investigation | Ari N. Machtinger, Validation, Writing-review &editing | Chris Crnich, Supervision, Project administration, Writing-review &editing | David H. O’Connor, Conceptualization, Funding acquisition, Writing-review &editing | Marc C. Johnson, Project administration, Funding acquisition, Resources, Writing-review &editing, | Shelby L. O’Connor Conceptualization, Data Curation, Formal analysis, Supervision, Visualization Validation, Project administration, Writing-original draft, Writing-review &editing.

## Data Availability

The authors confirm that data supporting the findings of this study are available within the article and its Supplementary data file. Raw data and metadata to perform these analyses are available in Supplementary Tables 7-11.

## Supplementary Material

Supplementary Table 1: Ct values derived from the GeneXpert instrument used in Figures 1-2 and Table 2.

Supplementary Table 2: Sampling period for all locations.

Supplementary Table 3: Total number of positives per virus on the GeneXpert System for all locations.

Supplementary Table 4: Results of statistical analyses comparing single virus application versus mixed virus application.

Supplementary Table 5: Results of statistical analyses comparing single virus application versus all virus application.

Supplementary Table 6: Results of statistical analyses comparing GeneXpert versus RT-qPCR.

Supplementary Table 7: Metadata of Wisconsin location A samples for heatmap generation.

Supplementary Table 8: Metadata of Wisconsin location B samples for heatmap generation.

Supplementary Table 9: Metadata of MU Student Center samples for heatmap generation.

Supplementary Table 10: Metadata of Columbia Boone County Courthouse samples for heatmap generation.

Supplementary Table 11: Data used for statistical analyses comparing GeneXpert versus RT-qPCR.

Supplementary Figure 1: ROC curves showing sensitivity-specificity trade-offs across GeneXpert Ct cutoffs.

