## Supplementary material for "Point-of-Care Air Surveillance of Respiratory Pathogens Using the GeneXpert® System": Supplementary Figure 1.docx

**Receiver operating characteristic (ROC) curves illustrating the trade-off between sensitivity and specificity across different classification thresholds for Cepheid GeneXpert**

**Figure 1:** The following figures show the ROC curve for 1) the SARS-CoV-2 **Missouri** data set, 2) the SARS-CoV-2 **Wisconsin** data set, and 3) the SARS-CoV-2 **Combined Missouri and Wisconsin** data sets, respectively.

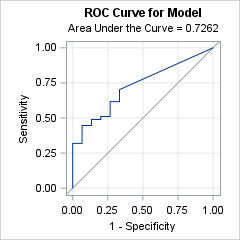

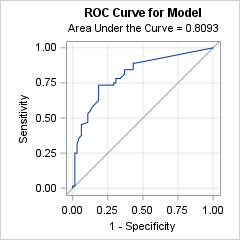

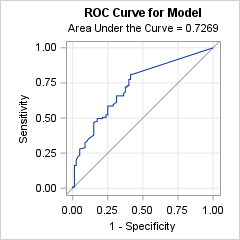

**Figure 2**: The following figures show the ROC curve for 1) **IAV A1** data set, and 2) **IAV A2** data set.

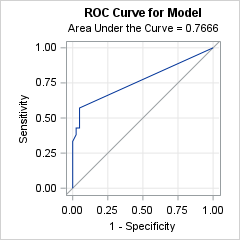

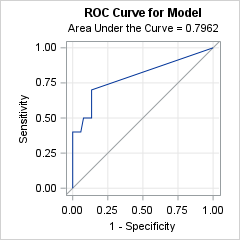
